# Impact of chlamydia and gonorrhea point-of-care testing on antibiotic prescribing in routine HIV care in rural Uganda

**DOI:** 10.64898/2026.04.22.26351517

**Authors:** Alex Abal, Julian Apako, Yobala B. Hurberd, Jacky Flipse, Guido J.H. Bastiaens, Erik Schaftenaar

**Affiliations:** Kumi Hospital, Kumi, Uganda; Department of Medical Microbiology, Makerere University, Kampala, Uganda; Department of Medical Microbiology, Rijnstate hospital, Arnhem, The Netherlands; Laboratory for Medical Microbiology and Immunology, Dicoon, Elst, the Netherlands; Stichting Global Health Initiative, Vianen, The Netherlands; Department of Medical Microbiology and Immunology, St. Antonius hospital, Nieuwegein/Utrecht, The Netherlands

**Author notes:** These authors contributed equally to this work (Alex Abal, Julian Apako).

## Abstract

**Objectives:** To evaluate whether on-site molecular point-of-care testing (POCT) for *Chlamydia trachomatis* (CT) and *Neisseria gonorrhoeae* (NG) is associated with reduced antibiotic overtreatment for presumed sexually transmitted infections (STIs) among adults living with HIV in rural Uganda.

**Methods:** We conducted a single-site quasi-experimental pre-post intervention study at Kumi Hospital, comparing syndromic management (April-August 2024) with CT/NG POCT-guided management (September 2024-January 2025). Adults living with HIV presenting with symptoms suggestive of an STI were included. Overtreatment in the pre-intervention phase was estimated by comparing antibiotic prescribing with the expected number of CT/NG infections based on positivity observed during the intervention phase.

**Results:** A total of 404 participants were included (203 pre-intervention, 201 intervention). During the intervention phase, CT and/or NG were detected in 14 individuals (7.0%). Median test turnaround time was 95 minutes, enabling same-day treatment in 93% of positive cases. Antibiotic prescribing decreased from 99.0% to 11.4% following POCT implementation (*P < 0*.*001*), corresponding to an absolute reduction of 87.6 percentage points. Estimated overtreatment declined from 30.0% to 5.0% for NG and from 74.9% to 6.0% for CT (both *P < 0*.*001*).

**Conclusions:** Implementation of CT/NG POCT in routine HIV care was associated with a marked reduction in antibiotic prescribing and estimated overtreatment for presumed STIs. These findings support the potential of POCT-guided, aetiology-based STI management to reduce unnecessary antimicrobial exposure in settings where syndromic management remains standard practice.

## INTRODUCTION

Sexually transmitted infections (STIs) remain a major global health challenge, disproportionately affecting low- and middle-income countries (LMICs), and continue to contribute to adverse reproductive and sexual health outcomes, including infertility, ectopic pregnancy, chronic pelvic pain, and increased risk of human immunodeficiency virus (HIV) acquisition and transmission [1,2]. In sub-Saharan Africa, clinical management of STIs continues to rely primarily on syndromic algorithms. Although operationally practical, these algorithms have poor diagnostic accuracy, especially for women, and fail to identify a substantial proportion of asymptomatic infections [3-5]. The limited specificity of syndromic management results in considerable overtreatment with broad-spectrum antibiotics. This is especially concerning for *Neisseria gonorrhoeae* (NG), a pathogen of major global concern because of its rapidly evolving antimicrobial resistance [3,4,6]. These shortcomings are exacerbated in people living with HIV, who are more vulnerable to STIs and suffer greater consequences when infections are missed or undertreated [7]. Strengthening local diagnostic capacity and transitioning STI care from syndromic to aetiology-based management are important to improve STI control and reduce unnecessary antimicrobial exposure [3,4,6].

In Uganda, national STI guidelines still rely on syndromic diagnosis and management, especially in rural settings, thus limiting surveillance, resistance monitoring, and the ability to deliver targeted/appropriate therapy [8,9]. Recent studies conducted in low-resource and in African settings show that implementation of *Chlamydia trachomatis* (CT) and NG point-of-care testing (POCT) within routine services is feasible and acceptable to patients and care providers [10-12]. Nonetheless, data on its influence on antibiotic prescribing practices in routine HIV care are still limited. This study assesses whether the introduction of CT/NG POCT reduces antibiotic overtreatment for presumed STIs.

## MATERIALS AND METHODS

### Setting and study population

We conducted a quasi-experimental intervention study at Kumi Hospital with a defined pre-intervention phase and intervention phase, to evaluate the effect of on-site molecular POCT for CT and NG on unnecessary antibiotic treatment among adults living with HIV presenting with symptoms of STIs. Kumi hospital, a 300-bed private, not-for-profit general hospital in Eastern Uganda, serves a predominantly rural catchment population of approximately four million people and provides a wide range of medical services, including comprehensive HIV care. The antiretroviral therapy (ART) clinic operates under Uganda’s national ART programme, and the hospital laboratory operates a GeneXpert platform (Cepheid, Sunnyvale, California, USA), on which CT/NG testing was implemented during the intervention phase.

Adults (≥18 years) living with HIV and attending the ART clinic with clinical signs or symptoms suggestive of a STI were eligible for inclusion. Exclusion criteria were active pregnancy, antibiotic treatment for an STI in the four weeks preceding inclusion, and (for the intervention phase) inability to provide consent. For the pre-intervention phase, anonymized data were retrospectively collected from routine ART clinic records on 4 October 2024 covering the period April-August 2024, using a standardized anonymized case report form (CRF). Information included demographic characteristics, HIV-related parameters (most recent viral load, ART status), reported STI-related symptoms, and prescribed treatments.

During the intervention phase (1 September 2024-31 January 2025), eligible participants were recruited consecutively during regular clinic hours. After written informed consent, participants were interviewed using the CRF. An urogenital swab was collected from each participant and tested on-site for CT and NG using the Xpert CT/NG assay according to the manufacturer’s instructions. Participants who tested positive received appropriate antibiotic treatment and counselling. Those testing negative were managed in accordance with the clinician’s judgement.

All CRFs were reviewed daily for completeness, double-entered, and stored in a secure, password-protected database. Data quality was ensured through routine field editing, cross-checking with source records, and correction of inconsistencies.

### Definitions

Treatment regimens were categorized into four groups based on antimicrobial activity: those including at least one agent active against NG (NG regimen); those including at least one agent active against CT (CT regimen); those including agents active against both pathogens (CT/NG regimen); and those lacking activity against either pathogen (other regimen). CT and NG regimens were based on the WHO treatment guidelines (S1 table) [13,14].

In the intervention phase, overtreatment was defined as receiving an NG- or CT-active agent in the absence of a positive test result. Because diagnostic testing was not performed during the preintervention phase, we estimated the expected number of NG- and CT-positive cases by extrapolating the positivity distribution from the intervention cohort. In the intervention phase, the prevalence of NG-only positive cases was 4.5%, 2.0% for CT-only positive cases and 0.5% for CT and NG positive cases. Including CT/NG co-infections as positive for both pathogens, this corresponded to an expected total of 10 NG-positive cases and 5 CT-positive cases among the 203 pre-intervention patients. Accordingly, overtreatment was defined as prescribing more than 10 NG-active regimens or more than 5 CT-active regimens.

### Outcome

The outcome was the reduction in overtreatment for CT and/or NG, assessed by comparing the proportion of symptomatic patients empirically treated under routine syndromic management in the pre-intervention phase with the proportion of patients with a positive CT and/or NG test receiving treatment in the intervention phase.

### Statistical analysis

Data were analysed using Stata version 17 (StataCorp, College Station, TX, USA). Descriptive statistics summarized participant demographics and clinical characteristics. Categorical variables were compared between phases using Pearson’s χ^2^ or Fisher’s exact tests, and continuous variables were analysed using Student’s *t*-test or the Wilcoxon rank-sum test, as appropriate. Logistic regression models were used to explore factors associated with overtreatment and confirmed CT/NG infection. Descriptive analyses were used to compare antibiotic prescribing patterns between study phases and between sexes.

## RESULTS

### Population characteristics

A total of 404 adults living with HIV were included in the study: 203 (50.2%) during the pre-intervention phase and 201 (49.8%) during the intervention phase (Table 1). The median age was 46 years (IQR 37-53), and the cohort included 322 women (79.7%). Demographic characteristics were broadly comparable between phases, with no significant differences in age distribution or viral suppression rates. However, the proportion of women was higher in the pre-intervention phase (181/203, 89.2%) compared with the intervention phase (141/201, 70.1%; *P < 0*.*001*), indicating a difference in sex distribution between study periods. Because STI epidemiology and symptom presentation may differ by sex, this variable was accounted for in multivariable analyses. All participants were receiving ART at the time of enrolment. Across the combined study population, the most frequently reported STI-related symptoms were lower abdominal pain (67.7%), painful micturition (43.4%), and genital or urethral discharge (37.0%), which ranked highest in frequency in both cohorts. However, participants with more than one STI-related symptom were more frequently observed in the pre-intervention phase (169/198, 85.4%) than in the intervention phase (66/164, 40.2%; *P* < 0.001).

**Table 1.** Comparison of baseline characteristics of study participants in the pre-intervention and intervention phases.

|  |  | Pre-intervention phase (n=203) | Intervention phase (n=201) | P value |
| --- | --- | --- | --- | --- |
| **Demographics** |  |  |  |  |
|  | Age in years | 46 (37-50) | 45 (38-54) | 0.303 |
|  | Gender (female) | 181 (89) | 141 (70) | <0.001 |
| **HIV status** |  |  |  |  |
|  | On ART | 203 (100) | 201 (100) | - |
|  | Most recent viral load<sup>a,b</sup> |  |  |  |
|  | <400 copies/mL | 173 (90) | 171 (88) | 0.57 |
|  | >400 copies/mL | 19 (10) | 23 (12) |  |
|  | **STI-related symptoms<sup>b</sup>** |  |  |  |
|  | Lower abdominal pain | 158 (80) | 87 (53) | <0.001 |
|  | Painful micturition | 97 (49) | 60 (37) | 0.013 |
|  | Genital/urethral discharge | 100 (51) | 34 (21) | <0.001 |

Data are shown as number (%) or median (interquartile range); ART, antiretroviral therapy; p value, Pearson Chi-square, Fisher's exact test or Mann-Whitney-Wilcoxon U test.
<sup>a</sup>p value was calculated for patients with suppressed viral load (<400 copies/mL). Missing data,
<sup>b</sup>Percentages are calculated using non-missing observations, missing data were excluded from the denominator.

### POCT test results

Among the 201 study participants in the intervention phase, on-site testing identified CT and/or NG infection in 14 individuals, corresponding to an overall prevalence of 7.0%. Nine (4.5%) were NG-positive only, 4 (2.0%) CT-positive only, and 1 (0.5%) positive for both pathogens. Positivity was slightly higher among females (7.8%) than males (5.0%), though this difference was not statistically significant (*P* = 0.56). All CT/NG Xpert assays were successfully completed, with a median turnaround time of approximately 95 minutes from specimen collection to result availability, enabling same-day treatment for 13 of 14 (92.9%) positive cases.

### Antibiotic prescription patterns

Participants in the pre-intervention phase were more likely to receive antibiotic treatment for presumed bacterial STIs [201/203 (99.0%)] than participants in the intervention phase [23/201 (11.4%), *P < 0*.*001*] (Fig 1). Antibiotic use was defined as the proportion of participants receiving at least one CT- or NG-active antibiotic regimen; therefore, the analysis reflects treated individuals rather than the total number of antibiotic doses administered. This corresponds to an absolute reduction of 87.6 percentage points in antibiotic use between phases.

**Fig 1.**
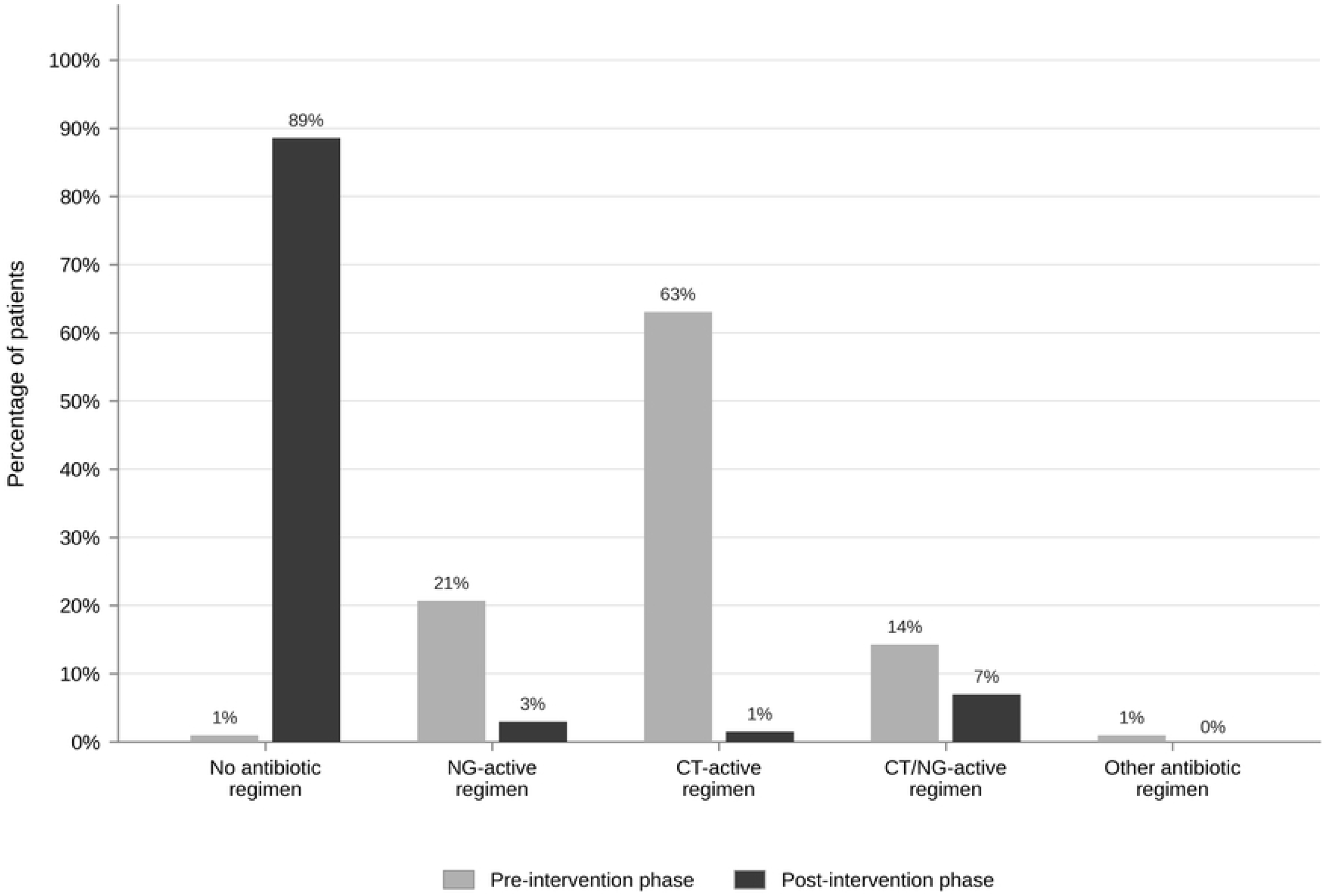
Treatment regimens in the pre-intervention- and intervention phases. NG, *Neisseria gonorrhoea*; CT, *Chlamydia trachomatis*.

In the pre-intervention phase, 128 participants (63.1%) received a CT-active regimen, 42 (20.7%) received an NG-active regimen, 29 (14.3%) received a combined CT/NG-active regimen, and 2 (1.0%) received an antibiotic regimen without activity against CT or NG. In contrast, in the intervention phase, only 3 participants (1.5%) received a CT-active regimen, 6 (3.0%) received an NG-active regimen, 14 (7.0%) received a combined CT/NG-active regimen, and none received an antibiotic regimen without activity against CT or NG.

In the intervention phase, all participants with a positive test result received an antibiotic regimen that appropriately covered the detected pathogen(s). Specifically, all 9 participants with NG-only infection (100%) received NG-active treatment, of whom 1 (11.1%) received an NG-only regimen and 8 (88.9%) received a combined CT/NG-active regimen. All 4 participants with CT-only infection (100%) received CT-active treatment; none received a CT-only regimen, whereas all 4 (100%) were treated with a combined CT/NG-active regimen. The single participant with dual CT/NG infection (100%) received a regimen active against both pathogens. Among the 187 test-negative participants, only 9 (4.8%) received an antibiotic regimen. Of these, 5 (55.6%) received an NG-active regimen, 3 (33.3%) received a CT-active regimen, and 1 (11.1%) received a combined CT/NG-active regimen, reflecting occasional empiric treatment despite negative test results.

### Overtreatment

Based on the prevalence observed in the intervention phase (4.5% NG-only, 2.0% CT-only, and 0.5% dual CT/NG infection), we extrapolated these proportions to the pre-intervention cohort, yielding an estimated 9 NG-only infections, 4 CT-only infections, and 1 dual infection among pre-intervention participants. Using these expected infection counts, study participants in the pre-intervention phase were more frequently overtreated for NG [61/203 (30.0%)] than participants in the intervention phase [10/201 (5.0%); *P* < 0.001]. A similar pattern was observed for CT, with overtreatment occurring in 152 of 203 participants (74.9%) in the pre-intervention phase compared with 12 of 201 participants (6.0%) in the intervention phase (*P* < 0.001).

## DISCUSSION

This quasi-experimental study among adults living with HIV attending a rural ART clinic in Eastern Uganda demonstrates that implementation of on-site molecular CT/NG point-of-care testing was associated with a substantial reduction in antibiotic prescribing for presumed STIs. Under routine syndromic management nearly all symptomatic patients received empirical antibiotic therapy, whereas following POCT implementation treatment was largely restricted to individuals with microbiologically confirmed infection. These findings highlight the considerable contribution of syndromic algorithms to unnecessary antimicrobial exposure in routine HIV care. While the direction of this effect is consistent with expectations, this study provides quantitative real-world evidence of the magnitude and operational feasibility of implementing aetiological STI diagnosis within routine HIV care in a rural sub-Saharan African setting. The median turnaround time of 95 minutes enabled same-day treatment for most patients with positive results, illustrating the operational feasibility of integrating molecular STI diagnostics into outpatient HIV services. The very low rate of antibiotic prescribing among test-negative participants further suggests strong clinician adherence to test results and acceptance of withholding empiric treatment.

Our findings provide quantitative evidence of the impact of CT/NG POCT on antibiotic prescribing practices in routine HIV care. The large reduction in antibiotic use following POCT implementation underscores the scale of overtreatment associated with syndromic management and highlights the potential of diagnostic testing to reduce unnecessary antimicrobial exposure, particularly in settings where concerns about antimicrobial resistance in NG are increasing. The CT/NG prevalence of 7% observed in the intervention cohort is consistent with estimates reported from comparable HIV care populations in sub-Saharan Africa [7,15]. In contrast, syndromic treatment algorithms implicitly assume a substantially higher prevalence among symptomatic patients.

Although the absolute number of antibiotic prescriptions declined substantially after POCT implementation, a relatively higher proportion of regimens included agents active against both CT and NG. This reflects guideline-based clinical practice, where dual-coverage therapy may be used when coinfection cannot be excluded or when clinicians treat confirmed infections with regimens covering both pathogens. In the intervention phase, most dual-active regimens were prescribed to individuals with microbiologically confirmed infection, suggesting that the observed pattern primarily reflects treatment decisions in a small number of confirmed cases rather than additional overtreatment.

This study has several limitations. First, the quasi-experimental pre-post design is susceptible to temporal trends and unmeasured confounding, and causal inference should therefore be made with caution. Second, differences in baseline characteristics between study phases, including sex distribution and symptom burden, may have influenced antibiotic prescribing patterns, although the magnitude of the observed reduction suggests that these factors are unlikely to fully explain the effect. Third, overtreatment in the pre-intervention phase was estimated by extrapolating CT/NG prevalence from the intervention cohort rather than directly measured, which may have resulted in misclassification if underlying prevalence differed between periods. Fourth, the study was conducted at a single rural HIV clinic, which may limit generalizability to other settings with different patient populations, healthcare structures, or diagnostic capacity. Finally, diagnostic testing was limited to CT and NG, and other causes of genital symptoms, such as *Trichomonas vaginalis* or bacterial vaginosis, were not assessed and may have contributed to residual antibiotic prescribing among test-negative individuals.

Cost and sustainability remain major challenges for wider implementation. Although the GeneXpert platform, already deployed in many LMIC settings for tuberculosis diagnostics, enabled rapid testing in this study, CT/NG cartridges remain expensive and require reliable electricity supply, maintenance, and procurement systems. These findings should therefore be interpreted primarily as proof of concept for diagnostic and antimicrobial stewardship rather than evidence supporting immediate large-scale implementation. Future work should include formal cost-effectiveness analyses, explore lower-cost molecular platforms, and evaluate targeted testing strategies focusing on higher-risk populations.

## CONCLUSIONS

In settings where syndromic STI management remains the standard of care, incorporation of molecular POCT into HIV care services could substantially reduce unnecessary antibiotic use and support more targeted STI management, provided that operational and economic barriers can be addressed.

## Data Availability

The minimal anonymized dataset underlying the results of this study is available upon reasonable request from the corresponding author. Due to ethical and legal restrictions related to the use of sensitive patient data, the dataset is not publicly available. Data access will be granted to researchers who provide a methodologically sound proposal and obtain appropriate ethical approval, where required. Requests may be directed to the corresponding author.

## ACKNOWLEDGEMENTS

The authors thank the Kumi Hospital staff for their support, collaboration and participation in this project.

## SUPPORTING INFORMATION

S1 Table. Treatment regimens based on the WHO guideline for the Treatment of *Neisseria gonorrhoeae* (2016) and WHO guideline for the Treatment of *Chlamydia trachomatis* (2016). NG, *Neisseria gonorrhoea*; CT, *Chlamydia trachomatis*.

